# Acute skin toxicity and cosmesis outcome in non-metastatic breast cancer patients treated with ultrahypofractionated radiotherapy: a randomized controlled phase II clinical trial comparing proton versus photon radiotherapy

**DOI:** 10.1101/2025.03.14.25323946

**Authors:** Thanaporn Sarsitthithum, Chonnipa Nantavithya, Chawalit Lertbutsayanukul, Kitwadee Saksornchai

## Abstract

**Background:** The 1-week photon radiotherapy schedule of 26 Gy in 5 fractions is as safe and effective as the standard 3-week schedule of 40 Gy in 15 fractions for adjuvant local radiotherapy in early-stage breast cancer. Proton radiotherapy offers dosimetric advantages by sparing the heart and lung dose but may cause more acute skin toxicities than conventional fraction photon radiotherapy. The acute toxicity of ultra-hypofractionated proton therapy is currently unknown.

**Materials and Methods:** In this trial, non-metastatic breast cancer patients were randomly assigned to receive either proton or photon therapy with the same dose of 26 Gy in 5 fractions given on alternate days. The aim of this trial was to compare acute skin toxicity between proton radiotherapy and photon beam radiotherapy using ultra-hypofractionation.

**Results:** Out of 140 eligible patients, 72 were included in this initial analysis (36 proton, 36 photon). Only one patient in the proton arm experienced grade 2 radiation dermatitis, which was the highest recorded grade. The incidence of acute radiation dermatitis was significantly higher in the proton arm compared to the photon arm (97.2% vs. 75%, respectively, p=0.006). There was no significant difference in other acute toxicities between the two arms. Patients treated with photon had better cosmetic outcomes (41.7% vs. 13.9%, P= 0.007) and were more satisfied (80.6% vs. 58.3%, P=0.04) than those in the proton arm. At a median follow-up of 9 months (IQR: 6-11), no locoregional recurrence was observed between the two arms.

**Conclusion:** In summary, this interim analysis found that ultra-hypofractionated proton therapy led to a higher rate of acute radiation dermatitis than the photon arm, although the dermatitis was mild. Patients in the photon arm had better cosmetic outcomes and higher satisfaction scores. However, further patient accrual and long-term follow-up are necessary to determine the long-term effects and effectiveness of proton therapy in breast cancer treatment.

## Introduction

Breast cancer is the most prevalent cancer in Thai women ^[1]^, and the standard treatment for early-stage breast cancer comprises surgery, chemotherapy, targeted therapy, and radiotherapy. Radiotherapy after breast-conserving surgery is crucial to reduce locoregional recurrence and breast cancer deaths ^[2, 3]^.

In King Chulalongkorn Memorial hospital, approximately 2000 new breast cancer patients have been treated at Division of Therapeutic Radiation and Oncology with conventional fractionation (45–50 Gy in 25 fractions of 1.8 or 2 Gy/day, 5 days a week, for 5 weeks) or hypofractionation (40.5-43.5 Gy in 15-fraction or 16-fraction of 2.65-2.7 Gy/day, 5 days a week, for 3 weeks) as recommended in international guidelines^[4, 5]^.

However, ultrahypofractionation, a 1-week schedule with a total dose of 26 Gy in 5 fractions, has demonstrated non-inferior tumor control^[6, 7]^ and provided several benefits in terms of patient convenience and cost reduction. Although concerns have been raised about late radiation toxicities due to the higher dose per fraction in ultrahypofractionation, the FAST-forward trial revealed that these toxicities were not inferior to those of hypofractionation^[7]^.

Due to the Bragg peak^[8]^, proton radiotherapy (PRT) has shown superior results in sparing heart and lung dose compared to photon therapy^[9-11]^. However, acute skin toxicities have been reported in early breast cancer patients receiving PRT through conventional fractionation^[12]^. When we compare the biological effective dose (BED) between conventional dose and ultrahypofractionated dose ^[7]^, the latter has lower value of BED (Appendix 1). Therefore, the acute responding tissue toxicity, i.e., skin, in ultrahypofractionated dose might not be as high as in conventional dose. If acute skin toxicity of ultrahypofractionated dose in PRT isn’t much higher than photon radiotherapy, using PRT will be beneficial for patients by sparing heart and lung dose, thus preventing late effects which can cause morbidity and mortality for patients.

Therefore, to our knowledge, this is the first randomized control trial aims to compare acute skin toxicity between PRT and photon beam radiotherapy using ultrahypofractionation in non-metastatic breast cancer patients.

## Methods

### Study design and patients

This is a single-center, non-blinded, phase 2 prospective randomized study conducted at King Chulalongkorn Memorial Hospital in Thailand. The eligible patients were women aged at least 21 years with pathologically confirmed breast cancer (stage 1-3 according to AJCC 8th edition), a good performance status (ECOG 0-2), and who underwent mastectomy or lumpectomy with negative margins. Axillary lymph node dissection or sentinel lymph node biopsy was performed according to the initial nodal status. Exclusion criteria included patients with active skin problems on the chest wall and axillary area, connective tissue disease, or other non-malignant systemic diseases that would prevent them from receiving study treatment or required follow-up, as well as pregnant or lactating women. Patients who had received radiation therapy for their currently diagnosed breast cancer prior to randomization, or prior radiation therapy to the ipsilateral chest wall or ipsilateral breast, as well as those who received bilateral whole breast or chest wall irradiation, were also excluded. The study protocol was approved by the Research Ethics Committee of the King Chulalongkorn Memorial Hospital (IRB number 017/64) and written informed consent was obtained from all patients. Patients who declined to take part in the study received the same standard of care as the patients who agreed to take part in the study.

### Randomization and masking

Patients were randomly assigned to receive either photon (control group) or proton therapy (intervention group) with the same dose of radiation by using the computer. Randomization was stratified by type of surgery (mastectomy or lumpectomy). Concealment was performed by a statistician and generated by the STATA program using the command “ralloc” for random allocation of treatment. Treatment allocation was not masked to clinicians or patients due to different radiation machines.

### Radiotherapy

This trial adopted a radiation dose of 26 Gy in 5 fractions from the FAST-Forward trial (5.2 Gy/fx) ^[7]^. Due to logistical constraints, we designed the radiation schedule to be given on alternate days. Patients with age <50 years or high grade (≥Grade 3) or close margins (<2mm) for both intervention and control groups received a tumor bed boost by using simultaneous integrated boost (SIB) techniques. This allowed physicists and physicians to use only one treatment plan. The SIB consisted of an additional 1 Gy/fx to the tumor bed with a total dose of 31 Gy for 5 fractions (6.2 Gy/fx). The SIB was given by proton in the experimental arm and photon in the control arm. In mastectomy patients who were randomized to the photon arm, a 1-cm bolus was added for 2 out of 5 fractions. To achieve a high skin dose in mastectomized patients in the proton arm, a 5-cm range shifter was applied in the treatment planning system.

For treatment planning, all patients were in supine position and immobilized using Vaclock/ Extend wing board (CIVCO MT 350) and both arms above the head. Computed tomography (CT) simulation was performed with a 3-mm slice thickness from the level of the larynx to the upper abdomen. CT simulation with either deep inspiration technique (DIBH) or free breath (FB) could be used according to the physician’s preference. CT images were transferred to Eclipse Planning System version 16.1 (Varian Medical system, Palo Alto, CA). The Clinical Target Volume (CTV) and Planning Target Volume (PTV) were delineated following the RADCOMP breast atlas v.3 guideline^[13]^ (Appendix 2). Daily positioning was achieved based on daily cone beam computed tomography (CBCT). The treatment plan was optimized to achieve the following PTV dose distribution and normal organ dose constraints as reference from our previous dosimetric study ^[14]^.

For proton arm, all the patients were treated with intensity modulated radiation therapy (IMPT) technique. The IMPT plans were generated with two fields: an anteroposterior (AP) field (0°) and an anterior oblique field with a gantry angle ranging from (+/-) 30° to 45° depending on the beam direction to be enface as possible to the breast or chest wall. Modifications to the beam dosimetric cover at all depths were made using a 5-cm range shifter. The Nonlinear Universal Proton Optimizer (V16.1) was used to optimize dose distribution. The Proton Convolution Superposition algorithm (V16.1) with a grid size of 2.5 mm and a constant relative biological effectiveness RBE of 1.1 was used to calculate the final dose. Robust optimization with a 5-mm setup uncertainty and a 3.5% range uncertainty was employed to take uncertainties into account and enhance target coverage and organs at risk sparing. All of the patients received treatment using a 6MV VMAT approach for photon planning. The right and left breasts’ VMAT plans included four arcs with gantry angles of 240° to 50° and 135° to 310°, respectively. Due to the limitation of the greatest left span of MLCs in the X-direction jaw of Varian linear accelerators, which is only 16 cm, the collimator rotated 90° for two arcs, splitting the jaw to cover the upper part and lower part of PTV, and increasing the coverage of dose in the PTV.^[15]^

### Assessments

All patients were evaluated and had physical examinations by researchers at the beginning and weekly during radiotherapy, then at 1st week, 2nd week, 1.5 months, and 3 months after completing radiotherapy. Subsequently, assessments were scheduled every 3 months until 1 year, every 6 months until 5 years, and then annually. Acute skin toxicities were graded using the standard CTCAE version 5.0 (Appendix 3) and were assessed weekly during radiotherapy, then at 1st week, 2nd week, 1.5 months, and 3 months after radiotherapy. To evaluate the cosmetic outcome of the breast and patient satisfaction, all patients were given a questionnaire with a four-point Harvard/NSABP/RTOG cosmesis criteria scale and a four-point Likert-type scale at the end of radiotherapy, at 1.5 months after radiotherapy, and at 3 months after radiotherapy (Appendix 4).

### Objectives

The primary outcome was acute skin toxicities focused on radiation dermatitis, defined as the worst grade that collected in each assessment within 3 months after radiotherapy. Secondary endpoints included locoregional recurrence (LRR), defined as recurrence of the disease at the ipsilateral breast or regional lymph nodes (supraclavicular, axilla and internal mammary lymph nodes), cosmetic outcome and satisfaction scores from patients and other toxicities such as skin infection, pain, pneumonitis, esophagitis and fatigue.

### Statistical analysis

The objective of this study was to compare acute skin toxicity rate between proton and photon beam radiotherapy with a different rate of 30% at a significance level of 0.05 and a power of 90%. It gave a calculation result of 112 in total sample size and plus 20% to drop out so, a total of 140 patients was needed. The interim analysis was planned when 50% of patients were included to investigate the toxicity of the method. The primary purpose of this analysis was to assess the toxicity profile of the treatment arms. The study also allows for early termination if a clear and clinically meaningful difference between the treatment arms is detected with sufficient statistical power.

The acute skin toxicities rate, good cosmetic outcome and satisfaction scores from patients were calculated by the binomial distribution and calculate the difference between proton beam radiotherapy and the photon beam with 95% confidence interval (95%CI) using two independent proportion Z-test. The difference in continuous data between two groups were calculated using an independent t-test. The difference in categorical data between groups were estimated using a Chi-square test or Fisher exact test. Statistical analysis was performed using STATA version 15.1. A P-value of less than .05 was considered statistically significant.

## Results

### Patient Population

Seventy-two patients (36 in the proton arm and 36 in the photon arm) out of a total of 140 patients who met the inclusion criteria were included in this interim analysis. Recruitment for this analysis began in August 2021, when the HRH Princess Maha Chakri Sirindhorn Proton Center at Chulalongkorn Hospital was officially opened, and the last patient was recruited in July 2022 (Figure 1). The baseline characteristics were well-balanced between the two groups, as shown in Table 1. The mean age was 55.3 years (range 33-90), and over half of the patients had undergone breast-conserving surgery (69.44% in the proton arm, 63.89% in the photon arm). Most patients received combination chemotherapy, both neoadjuvant therapy (22.22% in the proton arm, 13.89% in the photon arm) and adjuvant therapy (55.56% in the proton arm, 52.78% in the photon arm). Two clinical T4 patients in the photon arm received neoadjuvant chemotherapy. Approximately 50% of patients in both arms received a tumor bed boost with the SIB technique. Most of the patients received radiation to the chest wall/whole breast (CW/WB) with or without the supraclavicular (SPC) region. Radiotherapy treatment details (dosimetric evaluation) for each subgroup are presented in Appendices 8-11.

**Figure 1.**
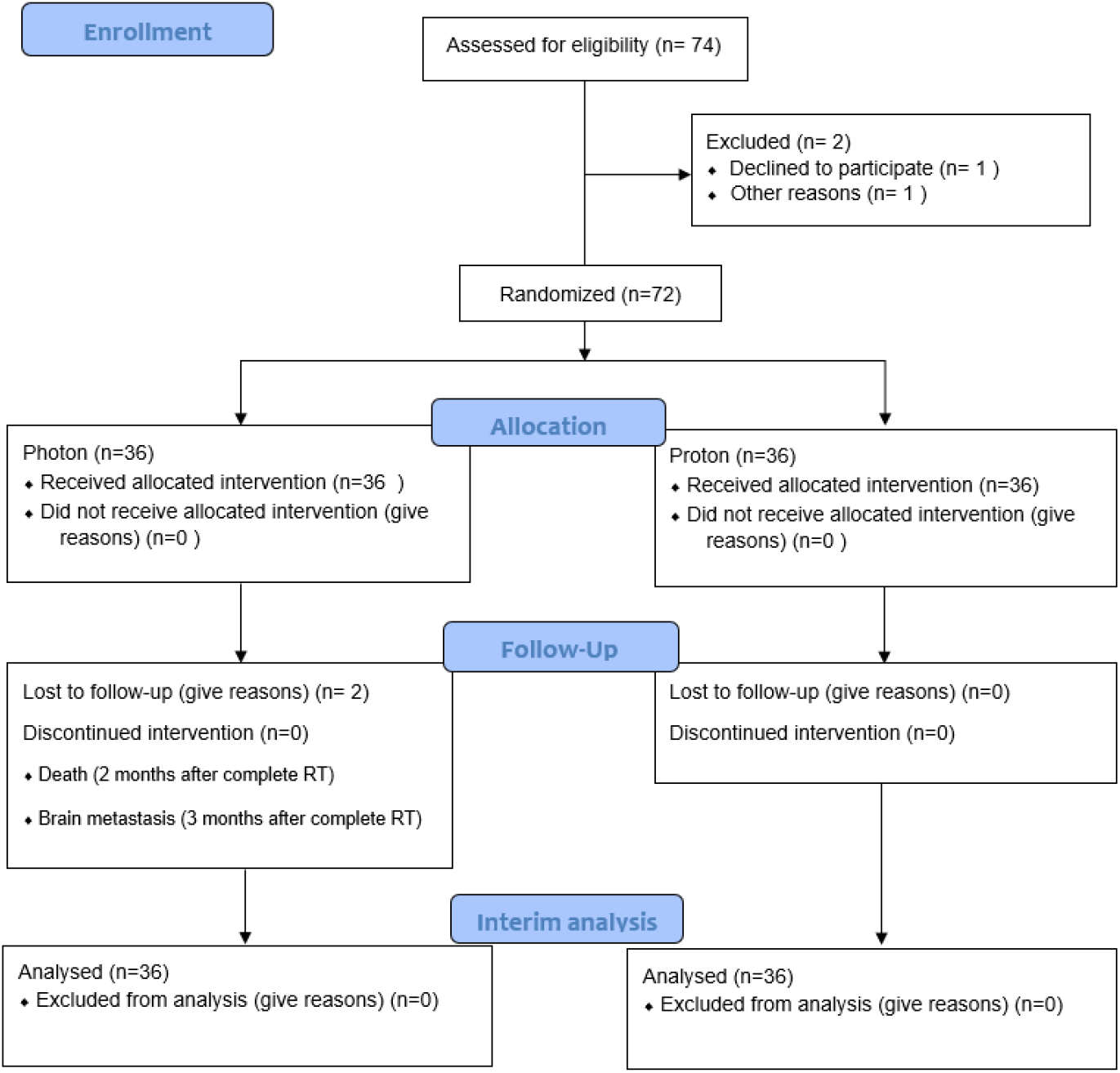
Consort flowchart.

### Acute toxicities

The toxicities of patients, as measured by the CTCAE v.5 during and after RT up to 3 months, were recorded as the highest grade. The primary outcome, the highest recorded grade of radiation dermatitis, was significantly higher in patients treated with protons (P = 0.01, Table 2). Only one patient in the proton arm had grade 2 dermatitis at 2 weeks after the complete RT session, which resolved to grade 1 skin dermatitis at the next follow-up. No patient experienced grade 3, 4, or 5 radiation dermatitis. Patients were categorized into groups that experienced radiation dermatitis toxicity versus no radiation dermatitis due to the absence of grade 3 and above toxicities. The incidence of acute radiation dermatitis was significantly higher in the proton arm versus the photon arm, showing a difference of 22.2% (P = 0.006, Table 3). Other acute toxicities were mild, with a range of gr 0 to 2, except for skin infection. One patient in the proton arm experienced grade 3 skin infection, which resolved after intravenous antibiotics. Other acute toxicities were mild and showed no significant difference between arms (Table 2). A univariate analysis found no significant association between dermatitis and factors such as SIB boost, radiotherapy volume, or type of breast surgery. (Table 4) In the proton arm, the dermatitis became more pronounced during the second week of radiation therapy and remained noticeable during the first 1-2 weeks post-radiation, after which it began to improve. In contrast, in the photon arm, dermatitis became more noticeable around the second week after starting radiation therapy. These patterns are clearly illustrated in the graph included in the Appendix5.

### Cosmetic outcome and Satisfaction scores

Details of the worst patient-rated outcomes and satisfaction scores are presented in Table 5. Patients treated with photon reported a higher proportion of excellent cosmetic outcomes (41.7% vs 13.9%, P= 0.007). Patients in photon arm rated “very satisfied” in a higher proportion than proton arm (80.6% vs 58.3%, P=0.04) (Table 5) A photo example from one patient who rated their cosmetic outcome as excellent is included in the Appendix6. Furthermore, as time passed, the cosmetic outcomes and satisfaction scores decreased, as illustrated in the graph provided in the Appendix7.

### Locoregional recurrence/ Distance metastasis

At a median follow-up of 9 months (IQR: 6-11), no locoregional recurrence was found in either arm. Two patients in the photon arm had distant metastases at 1 and 3 months, respectively. Both received mastectomy and systemic treatment from other hospitals before being referred for radiotherapy at our hospital. One patient in the photon arm had bone and liver metastases. She was initially classified as having clinical T4, N2, and M0 disease. After neoadjuvant chemotherapy and mastectomy, the pathological stage showed ypT4N2M0 with extra-nodal extension. Immunohistochemistry showed hormone sensitivity (ER-positive, PR-negative, HER2-negative). Liver and bone metastases were detected while she was receiving hormonal treatment at 1 month after completing radiotherapy. She passed away 2 months after completing radiotherapy. The other patient developed brain metastases 3 months after completing radiotherapy. She was initially classified as having clinical T3, N3, and M0 disease. Neoadjuvant chemotherapy followed by mastectomy was performed. Her pathological stage was ypT1N1M0 with HER2 overexpression. She is still alive and receiving systemic treatment from a medical oncologist. No distant metastasis failures were found in the proton arm.

## Discussion

Our previous dosimetric study demonstrated the dosimetric advantages of proton therapy in sparing the heart and lungs compared to photon therapy ^[14]^. This randomized controlled trial is the first to compare acute skin toxicity between proton radiotherapy and photon beam radiotherapy using ultrahypofractionation in non-metastatic breast cancer. In this interim analysis with 72 patients out of the total 140, mild (grade 1-2) radiation dermatitis was found to be significantly higher in the proton arm compared to the photon arm. However, it is important to highlight that no grade 3 dermatitis was observed in either arm, including the proton arm. Only one patient in the proton arm experienced grade 2 radiation dermatitis, while the rest of the cases were limited to grade 1 dermatitis, which is typically minimally symptomatic for patients. This suggests that proton therapy with ultrahypofractionation is feasible, resulting in minimal skin toxicity, especially when compared to earlier studies using more protracted fractionation regimens.

Similarly, patient-reported excellent cosmetic outcomes and a “very satisfied” satisfaction score were higher in the photon arm than in the proton arm. At a median follow-up of 9 months, no locoregional recurrence was found in either arm.

The mildness of the acute radiation dermatitis in the proton arm using the 5-fraction regimen (radiation dermatitis grade 1: 94.4%, grade 2: 2.8%) compared to the previously reported 25-fraction regimen ^[12]^ (radiation dermatitis grade 1: 30.8%, grade 2: 64.1%, grade 3: 5.1%) is likely due to the lower biological effective dose (BED) of ultrahypofractionation compared to conventional fractionation. Additionally, a higher proportion of mastectomies were performed in DeCecaris’s study compared to ours (mastectomy in proton arm: 53.8% vs. 30.56%).

Compared to the previous 5-fraction regimen in the FAST Forward trial ^[16]^ (radiation dermatitis grade 1: 58%, grade 2: 36%), our study showed a higher rate of grade 1 acute radiation dermatitis in the photon arm (radiation dermatitis grade 1: 75%) but a lower rate of grade 2 acute radiation dermatitis. This pattern also held true for the proton arm, which showed a notably low incidence of grade 2 dermatitis (3%) and no grade 3 dermatitis. The difference in treatment delivered as an everyday regimen in the FAST Forward trial (5 fractions per week) versus the every-other-day regimen in our study may have contributed to the reduction in acute toxicity in both arms.

A higher rate of acute radiation dermatitis in our study between the proton and photon arms, may be due to the difference in radiation technique in chest wall irradiation. In the proton arm, a 5-cm range shifter was applied in every fraction, while a 1-cm bolus was applied in only 2 fractions of treatment in the photon arm. Therefore, the skin dose in the proton arm was higher than in the photon arm in mastectomy patients. The only patient who experienced grade 2 radiation dermatitis and the only grade 3 skin infection in the proton arm was also a mastectomy patient. Due to this observation, caution should be applied in patients who receive chest wall irradiation. However, this is an interim analysis with only about 50% of expected accrual, of which only 11 mastectomy patients were in the proton arm.

Our study has some limitations. The total number of patients in this interim analysis is too small to interpret the difference in the primary outcome, especially in mastectomized patients, which had only 11 and 13 patients in the proton and photon arms, respectively. Second, we observed only mild acute radiation dermatitis occurred in both arms, and no locoregional recurrence was found. The other limitation was the short follow-up time of 9 months, so it cannot be concluded about locoregional control and late toxicities of a 5-fraction regimen, which needs further follow-up.

## Conclusion

This interim analysis demonstrated a significantly higher incidence of acute radiation dermatitis in the proton arm of non-metastatic breast cancer patients treated with an ultrahypofractionated dose. Although the observed radiation dermatitis was only mild (grade 1-2), patients in the photon arm reported superior cosmetic outcomes and higher satisfaction scores than those in the proton arm. Ultrahypofractionation with proton therapy, while providing normal organ sparing benefits, was associated with tolerable acute toxicities. Further patient accrual and long-term follow-up are required.

## Data Availability

All relevant data are within the manuscript and its Supporting Information files

